# A simple approach for multiple observations improves power to detect genetic effects and genomic prediction accuracy

**DOI:** 10.1101/2025.09.19.25336197

**Authors:** Luke M. Evans, Christopher H. Arehart, Raine A. Gibson, Grace I. Bowman, Christopher R. Gignoux

## Abstract

Many datasets, including widely used biobanks, have more than one observation of numerous phenotypes for at least a portion of their sample. The majority of GWAS utilize only a single observation per individual, even when more than one observation may be available, and apply a standard model in which the additive allelic effect being estimated is assumed to be constant across the age or time range in the sample. Here, we test a set of simple approaches to utilize multiple observations per individual, under this same assumption. We find that utilizing the mean or median of the available observations rather than a single observation improves power to detect associated loci and enriched gene sets and yields higher out-of-sample polygenic score prediction accuracy. Despite growing biobanks, many deeply phenotyped samples are relatively small but have multiple observations. While explicitly modeling age- or time-dependent genetic effects can estimate time- or age-specific genetic effects, most GWAS apply a standard, additive-only model; a simple approach of using the mean or median can improve power by reducing “noise” in the phenotype, utilize standard, optimized software, and be particularly impactful for smaller samples, including samples of diverse genetic ancestry currently existing in widely used biobanks.

## Introduction

The standard genome-wide association study (GWAS) model estimates allelic effects applied to a single observation for each individual^1^ using the model,

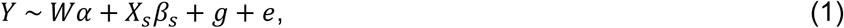

where *Y* is the vector of phenotype values, *W* is the matrix of fixed effect covariates, *α* is the vector of covariate effect sizes, *X_s_* is the genotype vector of all individuals at a particular single nucleotide polymorphism (SNP), *β_s_* is the per-allele effect of that SNP, *g* is a random effect that represents the contribution of other genetic variants, and *e* is the error, with *g* ∼N(0,*σ^2^_A_G*) and *e* ∼N(0,*σ^2^_E_*). *σ^2^_A_* is the additive genetic variance, *G* is the genetic relationship matrix, and *σ^2^_E_* is the error variance.

Many, perhaps most, large-scale GWAS apply this standard model (1), and span a range of phenotypes from height and body mass index to substance use and mental health, including large, recent studies^2–5^. For instance, two widely-used databases apply this standard model to 778^6^ or over 2000^7^ unique traits in the UK Biobank (UKB) dataset. The standard model (1) has fueled tremendous discoveries across many traits^8^ and is also appealing in that it is implemented in existing, optimized software^9,10^.

Embedded within this standard model (1) is a strong, yet frequently unstated assumption that the additive allelic effect is constant across the range of ages or time span of the sampled phenotypes. Age or study data collection wave is often included as a covariate in (1), accounting for mean differences in phenotype as a function of age or time^1^, but inclusion of covariates does not affect this strong assumption regarding the SNP effects themselves. With sufficient observations per individual, the time- or age-specific genetic effects can be modeled with specialized software packages^11–16^. However, many datasets have more than one observation for at least some individuals, but not enough to fully model the age- or time-dependent genetic effects. For example, the UK Biobank^17^ contains at least two observations for over 100,000 individuals for many phenotypes. While using specialized growth models or software^11–16^ may not be feasible with only two observation points, these additional observations are often ignored in favor of using either the data collection wave with the most observations or the most recent observation for each individual^2–7^.

Here, we explore whether these additional observations could be easily, realistically, and usefully incorporated into the framework of the standard GWAS through the use of commonly applied software packages to improve power to detect associations and predict phenotypes. Smaller datasets and lower-heritability traits may benefit the most from leveraging the power of multiple observations—in particular, datasets with very careful phenotyping over multiple waves of data collection. While often lost in larger, biobank-based GWASs, such datasets could still contribute greatly to our understanding of traits’ genetic etiologies. Further, strategies such as these are needed to improve power to detect genetic effects includes samples of diverse genetic ancestries. Most GWAS are performed using individuals of European genetic ancestry, in part because sample sizes are much smaller for samples of diverse genetic ancestry^18^.

An approach as simple as using a mean or median of all available observations could improve GWAS power, even when only a few observations per individual exist by reducing random noise in the phenotypic measurement. For instance, Chen et al.^19^ tested a range of approaches for leveraging multiple observations, and found that the median led to the highest estimated SNP-heritability. One possible consequence of this finding is that this approach could be implemented via the same software pipelines that are routinely used in genetic analyses, rather than requiring additional software packages or bespoke analytical pipelines. However, their analyses were restricted to only lipid traits in individuals of European genetic ancestry, did not assess GWAS power, and did not determine whether polygenic score prediction accuracy improves.

Here, we systematically test the incorporation of multiple observations using a range of different approaches across two well-characterized phenotypes – height and BMI. We focus on existing datasets—particularly those with relatively small samples of diverse genetic ancestry that are often removed for standard GWAS—to highlight how a simple approach applied to such samples can improve power to detect genetic loci, increase estimates of SNP-based heritability, and improve out-of-sample genetic prediction of these traits in samples of both European and diverse genetic ancestry. We demonstrate improved power and an up to 10% increase in out-of-sample polygenic prediction accuracy when using the mean of all available observations per individual instead of the most recent single observation, which arises because it reduces non-genetic, environmental noise of the phenotypic measurement. We encourage researchers to use this simple approach that can be readily implemented in standard pipelines to improve power and prediction accuracy in genetic studies.

## Material and methods

### Summary of methods

We first used simulated genotypes and longitudinal phenotypes to test whether different trait definitions that utilize multiple observations per individual impact a) the power to detect an associated locus, b) the accuracy of allelic effect size estimation, and c) the predictive accuracy of a polygenic score relative to the standard trait definition (the most recent observation per individual). Next, we applied multiple trait definitions to real longitudinal data from two independent datasets, performing GWAS, SNP-heritability, genetic correlation, and polygenic prediction analyses, comparing across different approaches to incorporate multiple observations. We applied these analyses separately to samples of European and diverse genetic ancestry to determine whether such simple applications of multiple observations can particularly improve detection of genetic associations and polygenic prediction in samples that are typically smaller, with a goal of addressing issues of equity in genomic medicine.

#### 1. Simulations

We simulated 20,000 individuals generating phenotypes under a polygenic model with 2,000 functional variants, varying the additive genetic variance for low heritability (*h^2^*=0.1) or moderate heritability (*h^2^*=0.5), and including gene-environment interaction (GxE) variance or not (*h^2^_GxE_*=0 or 0.05). We simulated phenotypes across 20 waves of data, varying whether or not there was a main, linear effect of wave and whether or not there was added environmental noise at later waves (heteroskedasticity with respect to wave). We further tested the effect of sample size (10,000 vs. 20,000 individuals) and including random missingness of 50% across waves. Using these simulated phenotypes, we then extracted the most recent observation, the mean of all observations, the median observation, and the integral of the fitted loess curve across all waves (see below for trait definitions). With each trait definition, we then estimated the additive effect of alleles under a standard GWAS model. For each parameter combination (varying *h^2^*, *h^2^_GxE_*, data collection wave, heteroskedasticity, N, missingness), we performed 1,000 simulations, recording the number of significantly associated loci, the correlation between estimated and true additive allelic effects, and the out-of-sample prediction accuracy for an identically generated, independent test sample.

All scripts to generate, analyze, and plot simulations are available on GitHub (https://github.com/evanslm/multi_obs_gwas), as are the numerical simulation results presented below.

#### 2. Phenotypes, Trait Definitions, and Datasets

We use the term “phenotype” to indicate the measured variable, such as height or BMI. We use the term “trait definition” to represent the operationalization of the measurement in the context of multiple observations per individual.

##### Trait Definitions

We applied seven different trait definitions to each phenotype: (1) mean of all available observations (R function *mean*), (2) median of all available observations (R function *median*), (3) maximum of all available observations (R function *max*), (4) most recent of all available observations, (5) trapezoidal integration of the area under all observations divided by the age range (R function *pracma::trapz*), (6) and the integration across age under the fitted loess curve (“loess”, R functions *loess* and *integrate*), and (7) estimated best linear unbiased predictors (BLUPs, the estimated subject-level random effect) using the *nlme* R package in a model that accounted for sex, *nlme::lme*(phenotype∼sex, random=∼1|subject). Trait definitions 1-2 captured the central tendency of an individual’s phenotype measurements, trait definition 3 captured the most extreme value, trait definition 4 represented single-point observations, trait definitions 5-6 summarized the longitudinal trajectories of observations (with and without smoothing), and trait definition 7 captured the subject-specific deviations from the fixed effect predictions. These seven different values for each subject were then used for each phenotype in GWAS or *h^2^_SNP_* analyses, as described below.

##### Datasets and Phenotypes Analyzed

We used the UK Biobank^17^ (UKB; application 1665) and the Health and Retirement Study^20^ (HRS; accessed via dbGaP accession phs000428, approved project ID 20774). Individuals within these datasets had between 1 and 15 observations across time (“waves”) for both height and BMI. The UK Biobank contains 1-5 observations per individual across time, spanning the in-person assessments and online follow ups, and self-report and linked health records. The HRS contains 1-15 observations per individual across 30 years of biennial data collection. Full descriptions of the specific variable and field IDs.

##### HRS

From the RAND 2020 HRS files, we extracted height from the RwHEIGHT and RwPMHEIGHT variables. There were 4,413 individuals of diverse genetic ancestry; 10,838 individuals of European genetic ancestry (EUR); and 10,776 unrelated individuals of EUR genetic ancestry (descriptions below). From the RAND 2020 HRS files, we extracted BMI from the RwBMI and RwPMBMI variables. There were 4,406 individuals of diverse genetic ancestry; 10,819 individuals of EUR genetic ancestry; and 10,759 unrelated individuals of EUR genetic ancestry (relatedness<0.05, used for GCTA *h^2^_SNP_* estimation [see below]). We also extracted age at each wave and self-reported sex for use as fixed effects covariates. We calculated mean, median, max, most recent, loess, trapezoidal, and linear mixed model random effect BLUPs for each phenotype. The HRS sample was used in the discovery sample for GWAS (see below) and *h^2^_SNP_* estimation.

##### UKB

For height (field ID 50), for individuals with ≥3 observations, 5,482 were of diverse genetic ancestry; 61,690 were of EUR genetic ancestry; and 47,526 were unrelated and of EUR genetic ancestry (relatedness<0.05, used for GCTA *h^2^_SNP_* estimation [see below]). For BMI (field ID 21001), for individuals with ≥3 observations, 5,610 were of diverse genetic ancestry; 67,037 were of EUR genetic ancestry; and 51,744 were unrelated and of EUR genetic ancestry. We also extracted age, sex, assessment center, and genotyping batch for use as fixed effects covariates. In the UKB, fewer observations per individual restricted the trait definitions we could apply, as there were not enough observations to calculate loess or trapezoidal trait definitions. For all individuals with ≥3 observations, we calculated mean, median, max, most recent, and linear model random effect BLUPs (“BLUPs”) for use in the discovery GWAS (see below) and *h^2^_SNP_* estimation. The remaining unrelated individuals, with 1 or 2 observations, were used as an independent testing sample for PGS analyses (see below).

##### Genetic Data Quality Control

We used array data for PCA and relatedness estimation, as well as for regenie^9^ whole-genome regression for GWAS (see below). We applied thresholds for quality control in plink^21^: biallelic, MAF>0.01, HWE p-value>1e-9, locus missingness<=0.05. For the UKB, we used the UKB-provided list of array markers they used for PCA. For HRS, we used HapMap3 positions and MAF- and LD-pruned array markers (plink --indep-pairwise 50 5 0.2). PCA was performed using FlashPCA^22^. Individuals of European genetic ancestry (EUR) were identified by projecting the sample onto the first four PC axes from 1000 Genomes^23^ (1KGv3) and identifying all individuals within +/- 5 standard deviations of the 1KGv3 EUR population mean, similar to other work^24,25^. The remaining individuals in each dataset represent individuals of diverse, non-EUR genetic ancestry, “DIV”. We estimated relatedness using GCTA^26^ for EUR genetic ancestry individuals or the GENESIS^27^ R package for DIV genetic ancestry individuals. Unrelated individuals (relatedness <0.05) were identified using GCTA.

We used imputed data for GWAS & SNP-heritability estimation. We used the UKB-provided HRC+1KGv3-imputed dataset, requiring biallelic sites, Imputation R2>=0.9, MAF>0.01, HWE p-value>1e-9, locus missingness<=0.05. For HRS, we used the Michigan Imputation Server for imputation to the Haplotype Reference Panel^28,29^, then applied the same thresholds. Thresholds were applied within subsets when appropriate, for example, in the slightly different sets of individuals with height vs. BMI data within HRS or UKB. QC thresholds were applied for each subset separately to ensure consistency.

#### 3. GWAS

In the European genetic ancestry samples, we used regenie^9^ to perform GWAS. We used the MAF- and LD-pruned genome-wide markers for the whole-genome regression. Covariates included the first ten principle component axes estimated within the EUR sample using flashpca and sex. For analyses of the UKBiobank, we also included Townsend Deprivation Index (field 189) and assessment centre (field 54) and age at time of first assessment (field 21003). For the HRS sample, we additionally included age as a covariate, with age measurement depending on the trait definition. For most recent observation, we included the age of that most recent observation; for maximum trait value, we included the age when that value was observed; for all others, we included the mean age of an individual. We performed GWAS using imputed genotypes for each phenotype and each trait definition. We meta-analyzed the effects of each trait definition for each phenotype across our two cohorts using inverse-variance weighted fixed effects meta-analysis in METAL^30^.

In the diverse ancestry samples, we used GENESIS^27^, following prior published work in pooled diverse samples^31^, as it better captures relatedness in the context of genetic structure within the sample. We first estimated PCs using PCRelate^32^, then estimated relatedness using those PCs using PCAir^33^. We then used GENESIS to perform linear mixed model GWAS using imputed genotype data, with the first five PCRelate PC axes, and other covariates as described above.

We applied standard follow-up approaches to interrogate the GWAS results. We first identified the number of conditionally independent loci using GCTA-COJO^34^, using the discovery GWAS genotype data for the LD reference and a p<5e-8 genome-wide significance threshold. We used GCTA-COJO for both the EUR-ancestry and diverse ancestry GWAS results. We then used DEPICT^35^ to identify enriched gene sets and tissues, and prioritized genes and loci. We applied DEPICT only to the EUR genetic ancestry samples as its distribution includes EUR genetic ancestry LD reference panels and used a p<5e-8 genome-wide significance threshold. We counted the number of significantly enriched or associated gene sets, tissues, genes and loci using FDR<0.05 to define significance, but also report counts based on strict Bonferroni correction.

Together, these analyses test whether different trait definitions have the same power to detect associations and gene set enrichment from GWAS. We hypothesized that using trait definitions that leverage multiple observations would reduce the non-genetic “noise” in the data, resulting in larger numbers of identified associations and enriched gene sets.

#### 4. SNP-heritability, Genetic Correlation, and Common Factor GSEM Modeling

We estimated SNP-heritability (*h^2^_SNP_*) and genetic correlation (*r_g_*) using the EUR-sample GWAS summary statistics within each cohort for each phenotype among all trait definitions. We used multivariate LD Score Regression (LDSC)^36^ implemented in the GenomicSEM^37^ R package, using the meta-analyzed summary statistics for each phenotype and trait definition, described above. We also estimated *h^2^_SNP_* and genetic and non-genetic variance components using GREML as implemented in GCTA^26,38^. We estimated GCTA-based *h^2^_SNP_* within each cohort separately, then used inverse variance weighting to meta-analyze the results.

We estimated common latent factor models for each phenotype using Genomic Structural Equation Modeling^37^ (GSEM), specifically to estimate standardized loadings of trait definitions for each phenotype within each cohort using GSEM. For a common factor model, standardized loadings near 1 and residual variances near 0 would indicate a latent factor that explains the shared variance in each indicator variable, while deviations from this would suggest unique genetic influences for certain trait definitions. Not all trait definitions were available across both cohorts; specifically, the trapezoidal sums and loess integration definitions were not estimated in the UKBiobank. Therefore, we used only mean, median, max, most recent and BLUP trait definitions. Because admixture impacts LDSC-based estimates of *h^2^_SNP_* and *r_g_^36,39^*, we performed these analyses only within the EUR samples.

Together, these analyses test whether different trait definitions are estimating the same genetic influences. We note several key features and specific hypotheses of these analyses. First, we note that heritability as a quantitative genetics concept refers to a specific population in a specific environment at a point in time^40,41^. While this idealized definition is difficult to meet in practice for any large biobank samples, our trait definitions specifically violate these assumptions – they are across time. We therefore use *h^2^_SNP_* as a useful *metric* to identify the *relative* strength of genetic and non-genetic influences, and do not interpret these estimates as heritability in the classic quantitative genetic definition sense. In this context, *h^2^_SNP_* is a useful scale to identify trait definitions that maximize the relative genetic contributions to the phenotype, i.e., those in which non-genetic “noise” has been removed, and therefore may have improved power to detect genetic effects. We therefore tested the null hypothesis that all trait definitions have the same estimated *h^2^_SNP_*, using clustered Wilcoxon rank sum tests (*clusrank* package^42^ in R), similar to other studies^19^. We also specifically tested whether the non-genetic variance (Ve) estimated from GREML were equal across trait definitions. In this vein, we then use genetic correlation estimates and common factor GSEM models to test whether the different trait definitions all share a common latent genetic structure. Even if one trait definition is noisier than another (and therefore presumably would have lower power to detect genetic associations and prediction accuracy in a PGS context), we still expect all trait definitions to have strong genetic correlations. We therefore tested the null hypotheses that all trait definitions of a phenotype within a cohort have *r_g_*=1 and that in a common factor GSEM model all have standardized loadings of 1.

#### 5. Polygenic Prediction Accuracy and Inference

We used LDpred2-auto^43^ with the LDpred2 LD reference panels and HapMap3+ set of markers, restricted to those sites for which our discovery GWAS included at least 20,000 individuals, as sites assayed in a single cohort (generally, the smaller HRS) and had much larger SE(beta) estimates which led to larger differences between the genotypic standard deviation estimated from the summary statistics compared to that estimated from the reference panel (see ref.^43^ for further details).

We next generated PGSs for each trait definition for each phenotype in the held-out individuals of the UK Biobank for individuals of European genetic ancestry, using either the most recent observation or the midpoint of two observations when present of individuals for each phenotype for unrelated individuals (those with 3+ observations were part of the discovery sample GWAS). We ensured no relatedness between the discovery UK Biobank sample and testing sample by estimating relatedness using GCTA^26^ across all discovery + test individuals and removing testing individuals as needed to ensure no pairwise relatedness with any discovery individual was >0.05.

To test the effect of the PGSs, we used a linear model that included the same covariates as before (see above), with or without the PGS. We estimated the incremental *R^2^*, or the adjusted *R^2^* difference between a full model (with PGS) to a reduced model (covariates only), as our metric of prediction accuracy. We estimated 95% confidence intervals by bootstrapping over individuals in the test dataset.

These analyses test whether different trait definitions have the same out-of-sample PGS predictive ability. We hypothesized that using trait definitions derived from multiple observations would reduce the non-genetic “noise” in the data and therefore have higher prediction accuracy.

## RESULTS

### 1. Simulations

In simulations, we found increased power, effect size estimation accuracy, and out-of-sample prediction accuracy when using multiple observations per individual, relative to only using the most recent, single observation in discovery GWAS (**Fig. 1, Table S1**). These effects were consistent across low (0.1) and moderate (0.5) heritability. When there were true GxAge effects, meaning the discovery GWAS model was misspecified because it only estimated the additive allelic effect, we still found improved power, accuracy, and prediction accuracy when using multiple observations. Notably, simulating true gene-by-age interaction effects resulted in strongly decreased correlation of true and estimated additive allelic effects when using a single observation, but not when using multiple observations (**Fig. 1**).

**Figure 1.**
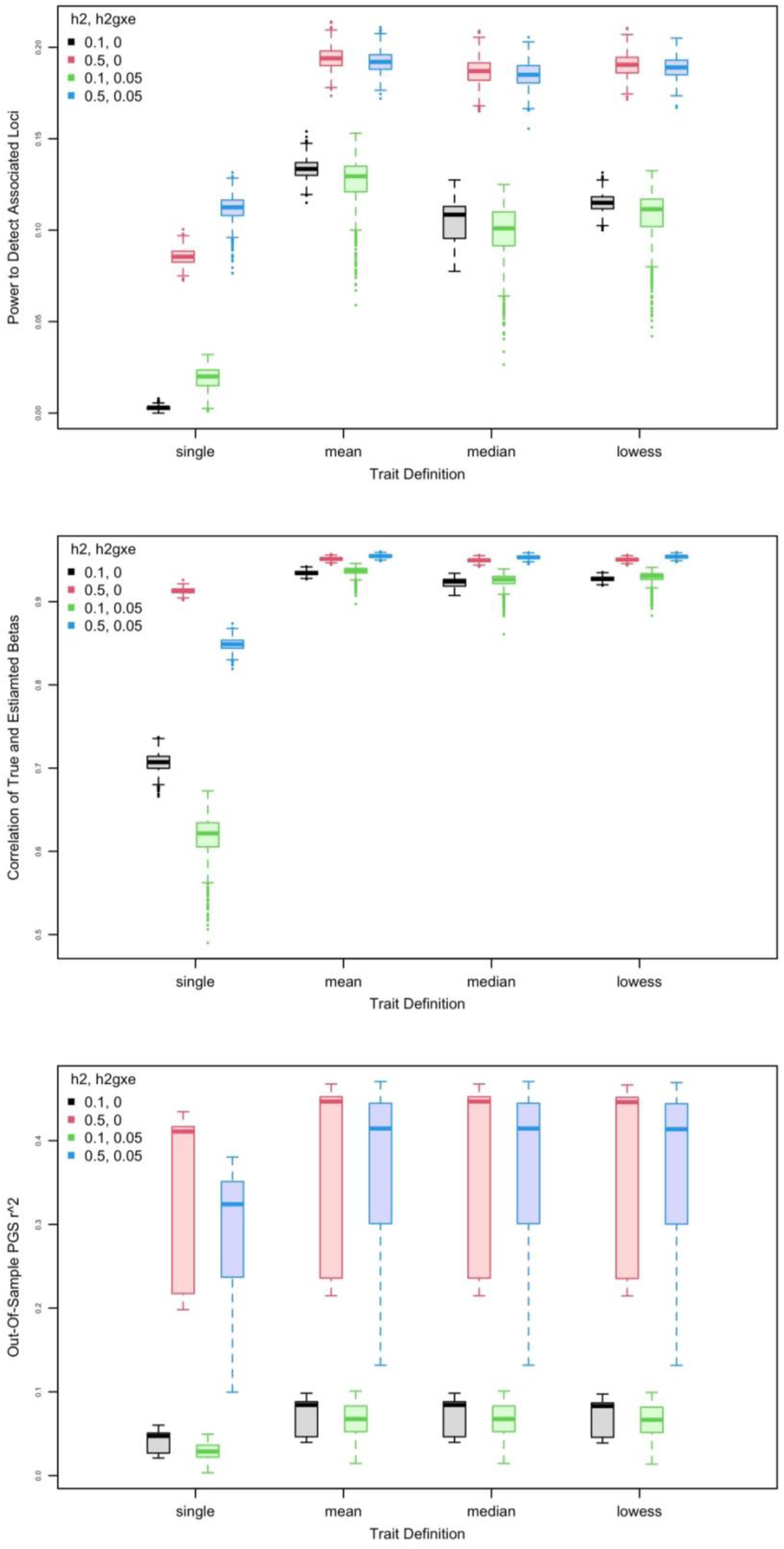
Simulation results estimating power to detect true associations (top), the correlation of true vs. estimated additive allelic effect sizes (middle), and out-of-sample prediction accuracy (bottom) when using different trait definitions. Colors represent the true overall additive genetic contribution (*h^2^*= 0.1 or 0.5) and the presence or absence of GxE (here GxAge) effects (h^2^gxe = 0 or 0.05). Note that in estimating the effects, only an additive model was used, replicating a standard GWAS.

Simulating a main, additive effect of age minimally impacted power to detect associations and the correlation of true and estimated allelic effects, but had strong, negative effects on out-of-sample prediction accuracy at high levels in particular (**Figs. S1-S3, Table S1**). Conversely, adding time-dependent heteroskedasticity had no significant effects on any of these outcomes (**Figs. S1-S3, Table S1**). Increased sample size had predictable effects, but the impact was greatest when only a single observation was used (**Fig. S4**). Random data missingness in the context of multiple observations had minimal effects (**Fig. S5**).

### 2. GWAS

Using GCTA-COJO, we identified consistently fewer conditionally independent significant associations using the single, most recent observation for all phenotypes when using the larger EUR discovery sample (**Fig. 2**). Overall, the mean, median, max and subject random effect trait definitions were all very similar (**Table S2, S3**). In the diverse genetic ancestry sample, we found fewer significant associations overall (consistent with smaller sample size) but found more when using trait definitions based on multiple observations compared to a single observation (**Fig. 2, Table S2, S3**).

**Figure 2.**
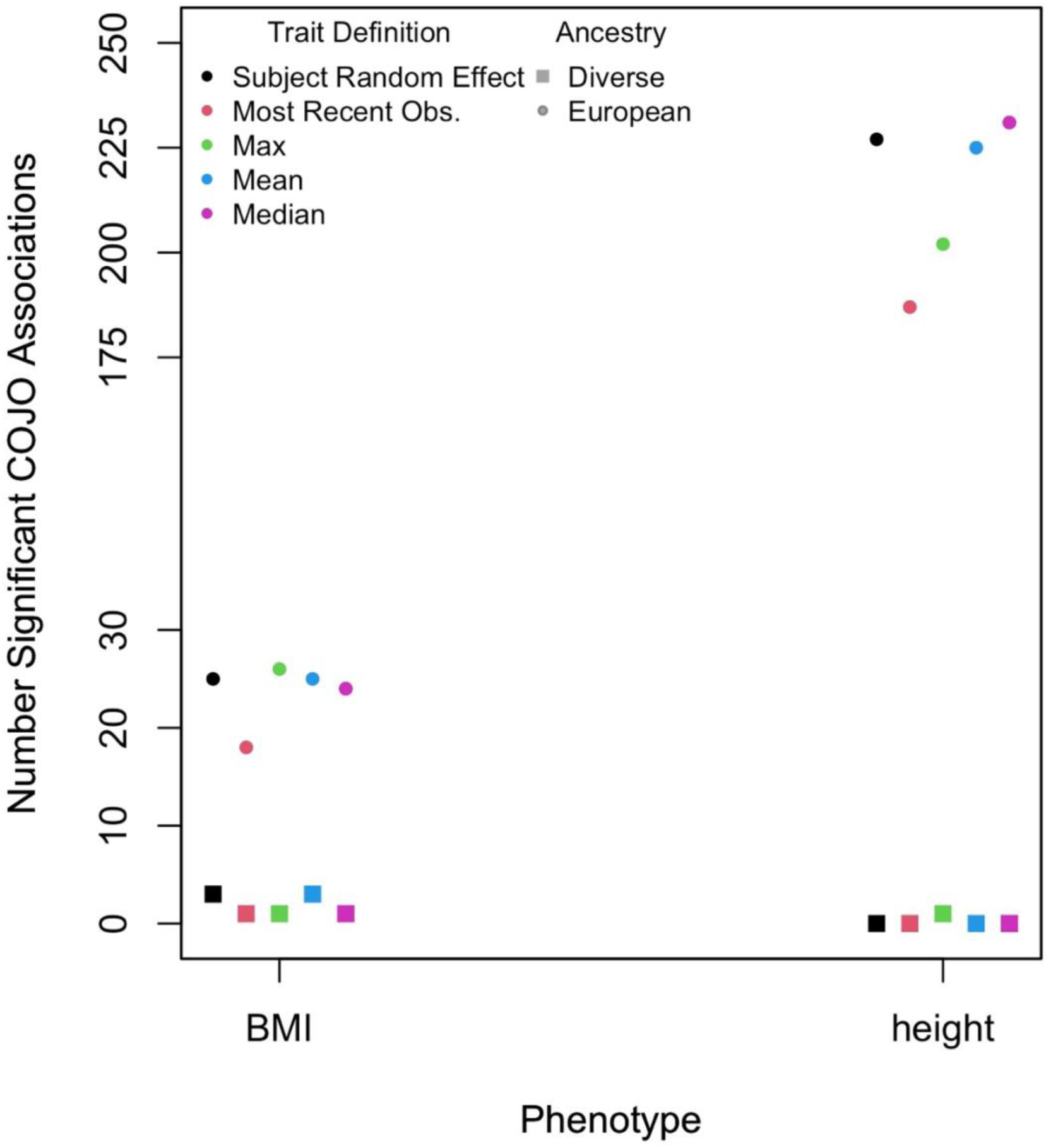
The number of conditionally independent significant associations for each phenotype using each trait definition for each discovery sample (p<5e-8). Note the break in the y-axis.

Using DEPICT for height, we typically found the fewest number of prioritized genes for significant loci, enrichment of gene sets, and tissue enrichment associations when using the single observation trait definitions (most recent or the maximum); other trait definitions using multiple time points produced largely similar results with more numerous prioritized genes, tissues, or gene sets (**Table S4**). For BMI, we found one significantly enriched tissue when using the median or max trait definition, and the fewest significantly associated loci when using a single most recent observation (**Table S4**).

### 3. SNP-Heritability, Genetic Correlation, and Common Factor GSEM

Across both phenotypes, using the most recent observation led to the lowest point estimate of SNP heritability using both GCTA and LDSC (**Figure 3**, **Tables S5-S6**). Using Wilcoxon signed rank tests with clustered data for each phenotype, all were nominally significant (0.028<p<0.044; **Table S5**). Examining the non-genetic variance (VE) estimated in GCTA within cohorts (**Table S5**), estimated VE was lower for the mean, median, max, and BLUPs trait definitions compared to when using the most recent observation.

**Figure 3.**
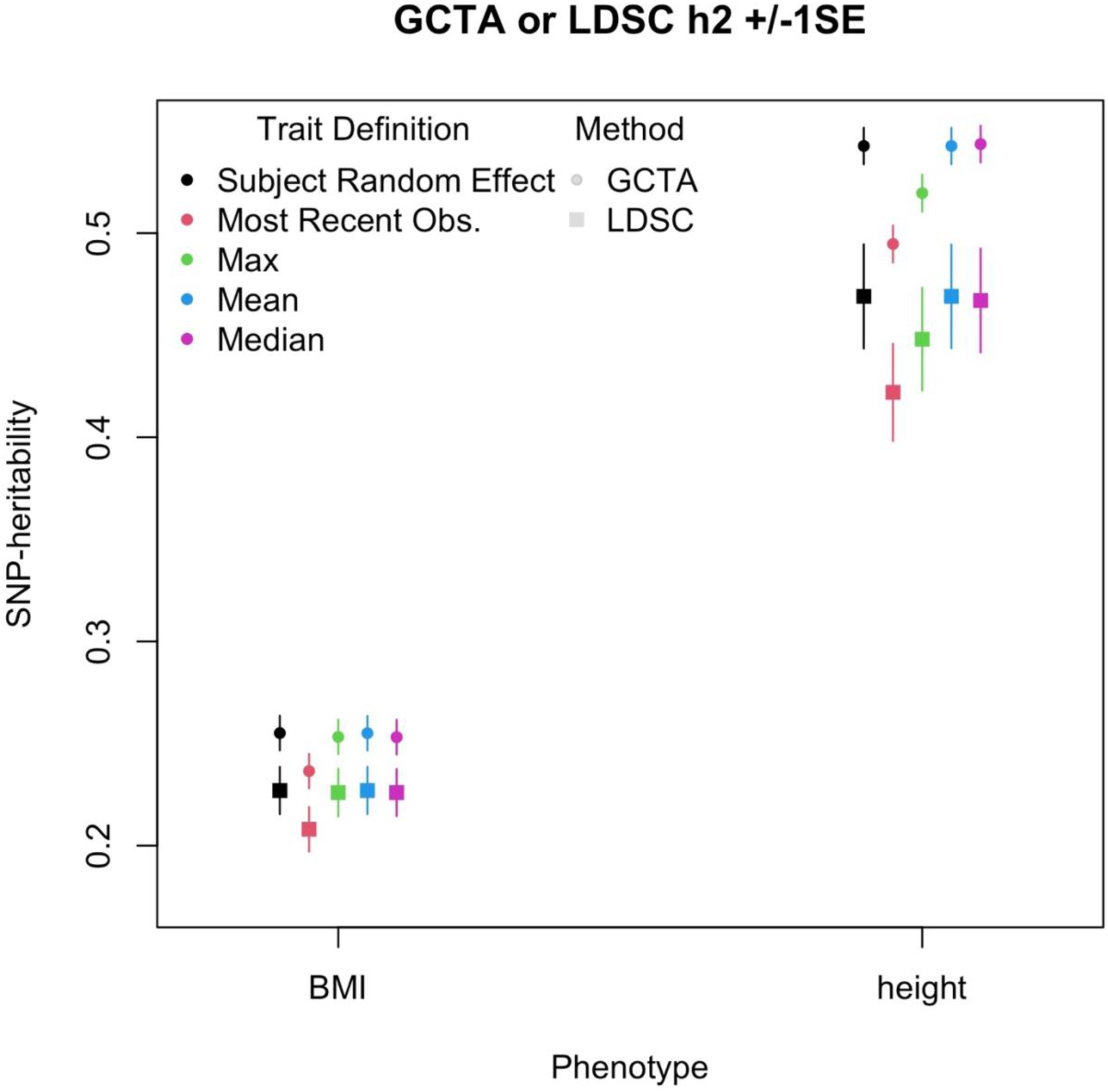
Estimated SNP-heritability +/- SE using either GCTA (circles) or LDSC (squares) for each phenotype (x-axis) and trait definition (colors).

While using the most recent observation led to lower SNP-heritability estimates, all trait definitions tapped into the same genetic phenotype, as indicated by nearly all 95% confidence intervals of pairwise genetic correlations among trait definitions for the same phenotypes not distinguishable from 1 (**Table S6**). This suggests all trait definitions are capturing the same genetic etiology for a given phenotype. Furthermore, no estimates of standardized factor loadings using a common factor model in GSEM were statistically different from 1 (**Table S6**).

Together, these empirical patterns recapitulate the simulation results, suggesting that using multiple observations can reduce non-genetic noise, leading to stronger genetic relative to non-genetic signal, which is the primary focus of standard GWAS.

### 4. Polygenic Prediction Accuracy

For all phenotypes, there was a significant effect of the PGS (all p<10^-14^) in the unrelated test dataset. Out-of-sample prediction accuracy of the PGS was up to 10% higher when using mean, median or subject random effects (subject BLUPs) relative to the most recent for BMI and height (**Fig. 4, Table S7**). For BMI, incremental *R^2^* when using the most recent observation was 0.0528, but 0.0583 when using the subject random effects, a 10.4% gain in PGS accuracy for the same sample size. Bootstrapped 95% CIs of *R^2^* were non-overlapping, indicating significant improvement of using multiple observations in the discovery GWAS compared to only the most recent observation. Similarly, for height, incremental *R^2^* using the simple mean was 0.114, but 0.109 when using the most recent observation, a 4.8% gain in PGS accuracy. Bootstrapped 95% CIs of *R^2^* were non-overlapping between using the most recent observation and using the median, max or subject random effect; 95% CIs narrowly overlapped when using the mean.

**Figure 4.**
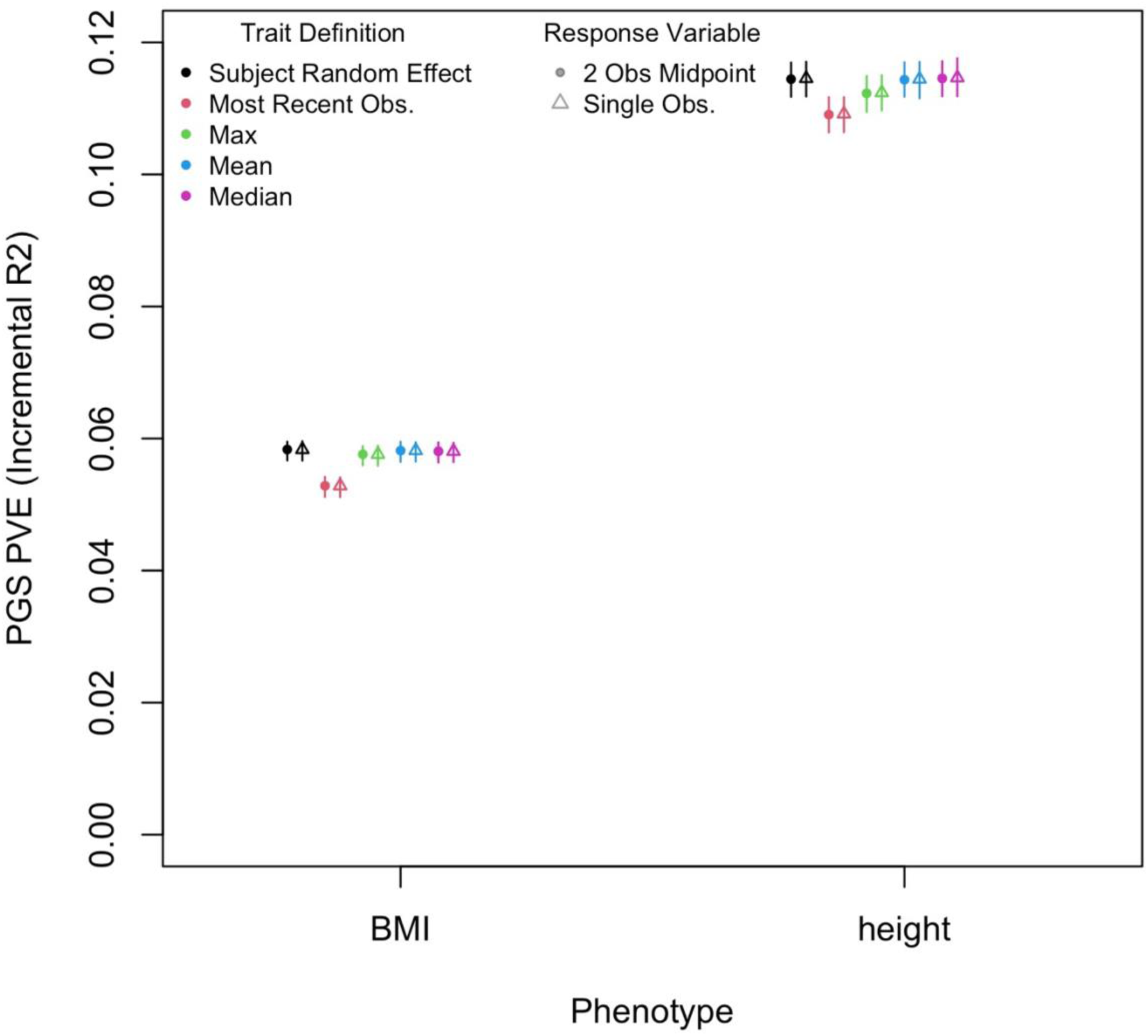
Out of sample prediction accuracy, as measured by the proportion of variance explained (PVE, or the incremental *R^2^* when the PGS is included relative to the reduced model) of the PGS for each phenotype and each trait definition, when using either the midpoint of two observations per individual or the most recent observation. Bars represent the bootstrapped 95% confidence interval from 1,000 bootstrap replicates.

## DISCUSSION

Most GWASs use a single observation of each individual, applying age or other time-related variables as linear, additive covariate effects. This carries a strong assumption that the genetic effect of an allele is constant across the range of ages in the study. Under this same assumption, we have shown that a simple, easy-to-implement approach—using the mean or median of each individual’s observations—improves power to detect loci, provides more accurate estimates of the additive allelic effect, and increases predictive accuracy of resulting polygenic predictions. Simply reducing non-genetic ‘noise’ in the phenotypic data improves the estimates from a GWAS, a well-known statistical point but one not typically implemented in the majority of GWASs, even when more than a single observation per individual is available.

When a research question is explicitly interested in age-related traits, modeling the phenotype using growth models that fit the *a priori* hypothesized functional form would have clear benefits, and our analyses and conclusions do not suggest otherwise. A productive future direction would be to characterize how using multiple observations would impact genetic estimates for such age-related traits, when not enough observations per individual exist for fully modeling those time- or age-dependent relationships.

We emphasize that this is a simple way that *existing* datasets can be used more productively. All GWAS perform some type of quality control on the phenotypic data. When even two or three observations are available for even some individuals in a dataset, a simple mean or median of those values de-noises the phenotypic data and improves GWAS precision and power. This is consistent with the existing, standard assumption when estimating a time-invariant additive genetic effect applied (though rarely explicated) in most GWASs, and also matches the theoretical definition of the additive effect of an allele substitution (sensu Falconer & Mackay^41^). Such an approach is easy to implement, in part because the resulting de-noised phenotypes can be directly used in widely used and optimized software (e.g., regenie^9^, GENESIS^27^, fastGWA^44^, Bolt-LMM^45^). While others have proposed modeling longitudinal data in a mixed-model framework^11–16^, which has additional benefits, they require more observations to estimate additional terms in the model (e.g., additive effects on the variance per trajGWAS^12^), and use separate software packages that require implementing another step in the process. This has additional benefits in terms of additional aspects of phenotypes analyzed but is a barrier to their use.

The benefits of this simple approach are particularly advantageous for smaller samples and lower-heritability traits when there are few observations, as seen in Fig. 1. Large biobanks linked to EHR data may provide longitudinal observations, but using EHR data has challenges on its own^46^, and the phenotypes are often broad phecode categories rather than specific and detailed. Alternatively, many smaller studies used in consortia may have more detailed phenotypes observed across more than one wave of data collection. Those detailed phenotypes may still be more productively used by simple de-noising strategies.

Smaller sample sizes are also characteristic of existing samples containing underrepresented ancestry groups in genetic studies. Often researchers remove these individuals from larger datasets^47^, yet we emphasize that such individuals can and should be used, and a simple de-noising of the phenotypes can improve power to detect genetic associations in smaller subsets of data. By simply using the means of individual observations of individuals of diverse genetic ancestry in one of the most widely used datasets (UK Biobank), we identified more significant BMI associations relative than when using a single observation per individual. This represents a tangible gain in equity, utilizing existing datasets and resources. This does not subvert the need for new, large and more ancestrally diverse samples; it is one of several strategies that will collectively improve equity in genomic medicine, and using all available approaches (and existing datasets) will provide the largest gains.

Limitations of our work include the relatively small sample sizes; the largest GWASs now contain 5M+ individuals^2^, and when scaled to these sizes, gains from using multiple observations may be limited. However, for most phenotypes, such large sample sizes are still out of reach, particularly for samples of diverse, non-European genetic ancestry. We have not evaluated how to consider binary traits or disease outcomes, but we note that random effects models such as those employed here can accommodate binary or other non-normally distributed data. Costs associated with collecting multiple observations can be significant, but longitudinal data has benefits beyond our main point of increased power in standard GWAS, and modeling gene-by-age or -time has potential to contribute to our understanding of age-related diseases and traits beyond the simple additive models we apply here.

Using simulations alongside analyses of power and predictive accuracy in both European and diverse genetic ancestry samples, we argue that an easy-to-implement approach – using the mean or median of all observations per individual in a dataset – can provide tangible gains in existing data and software under standard assumptions.

## Data and Code Availability

This study used data available through the UK Biobank and Health & Retirement Study, which can be accessed through those repositories. All scripts to generate, analyze, and plot simulations are available on GitHub (https://github.com/evanslm/multi_obs_gwas), as are the numerical simulation results presented below.

## Acknowledgements

We thank the participants of the UK Biobank and HRS, and we thank the studies and their administrators. This research has been conducted using the UK Biobank Resource (application number 1665). The HRS (Health and Retirement Study) is sponsored by the National Institute on Aging (grant number NIA U01AG009740) and is conducted by the University of Michigan. HRS data was accessed through dbGaP phs000428.

Data storage for this project was supported by the PetaLibrary and computational analysis was supported by the Blanca and Alpine high performance computing resources at the University of Colorado Boulder (funded by the University of Colorado Boulder, the University of Colorado Anschutz, and Colorado State University).

CHA was supported by the IBG NIMH T32MH016880 training grant and by the Interdisciplinary Quantitative Biology program. RAG was supported by the University of Colorado Boulder’s Summer Multicultural Access to Research Training program, part of the Colorado Diversity Initiative, which is funded internally by the University of Colorado Boulder Graduate School. GIB was supported by the Interdisciplinary Quantitative Biology Program and Hevolution/AFAR HEV∼NI23013. LME was supported by Hevolution/AFAR HEV∼NI23013 and NIA R01AG046938. We thank John Hewitt and Chandra Reynolds for helpful discussion.

