## Supplemental Figure for "A simple approach for multiple observations improves power to detect genetic effects and genomic prediction accuracy"

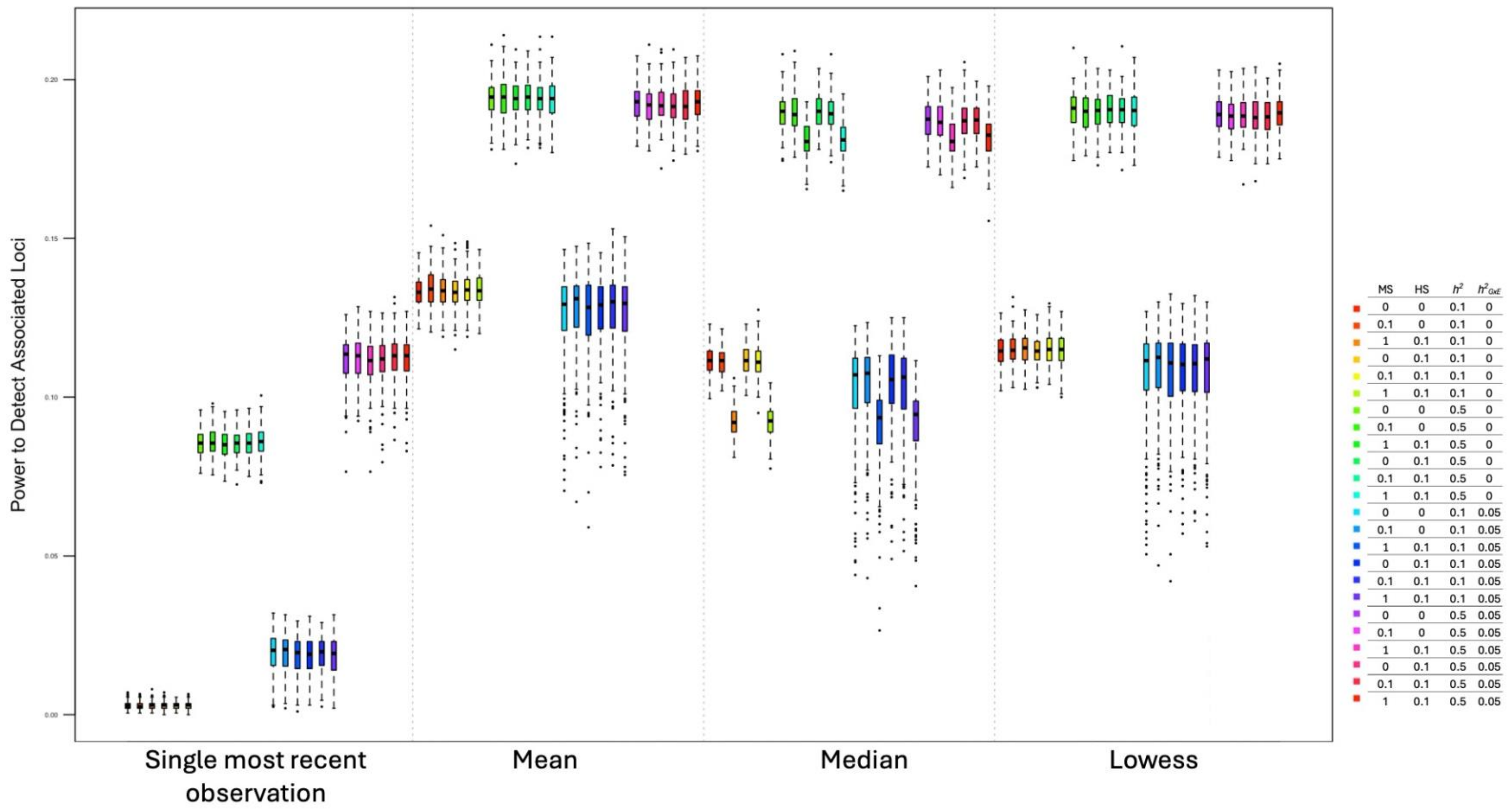


**Figure S1.** Power to detect truly associated loci using simulations with different parameter combinations (shaded colors). MS=main, additive slope of time, i.e., an increase in the overall trait value as a function of time. HS=time-dependent slope of heteroskedasticity, or the increased phenotypic variance as a function of time. $h^2$=simulated heritability. $h^2_{GxE}$=simulated gene-by-time variance as a proportion of overall phenotypic variance.

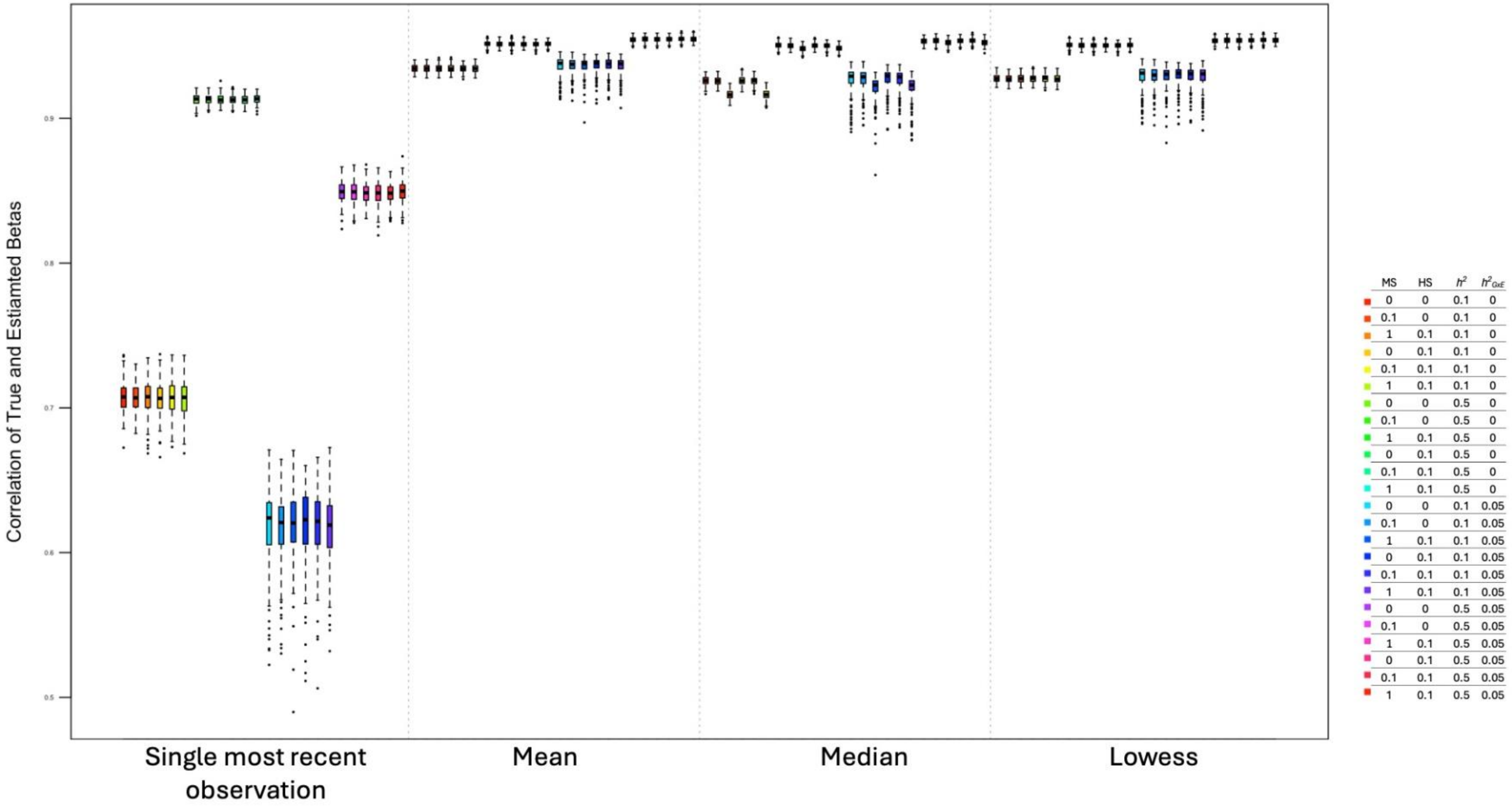


**Figure S2.** Correlation of true (simulated) allelic effects and estimated effects using simulations with different parameter combinations (shaded colors). MS=main, additive slope of time, i.e., an increase in the overall trait value as a function of time. HS=time-dependent slope of heteroskedasticity, or the increased phenotypic variance as a function of time. $h^2$=simulated heritability. $h^2_{GxE}$=simulated gene-by-time variance as a proportion of overall phenotypic variance.

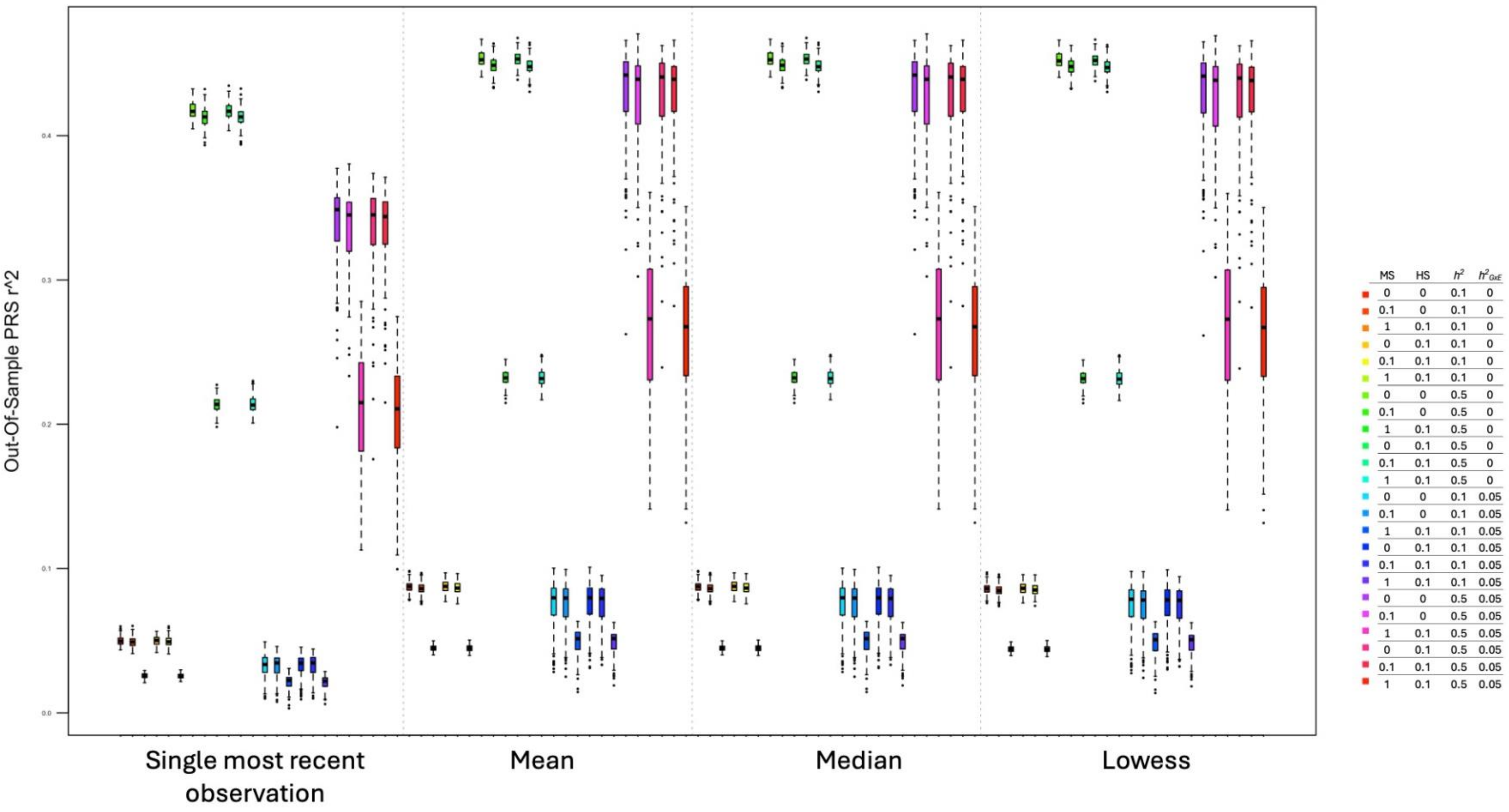


**Figure S3.** Our-of-sample prediction r2, measured as the squared correlation between true simulated phenotypic values and the PRS-predicted phenotype, using independent discovery GWAS and target samples, using simulations with different parameter combinations (shaded colors). MS=main, additive slope of time, i.e., an increase in the overall trait value as a function of time. HS=time-dependent slope of heteroskedasticity, or the increased phenotypic variance as a function of time. $h^2$=simulated heritability. $h^2_{GxE}$=simulated gene-by-time variance as a proportion of overall phenotypic variance.

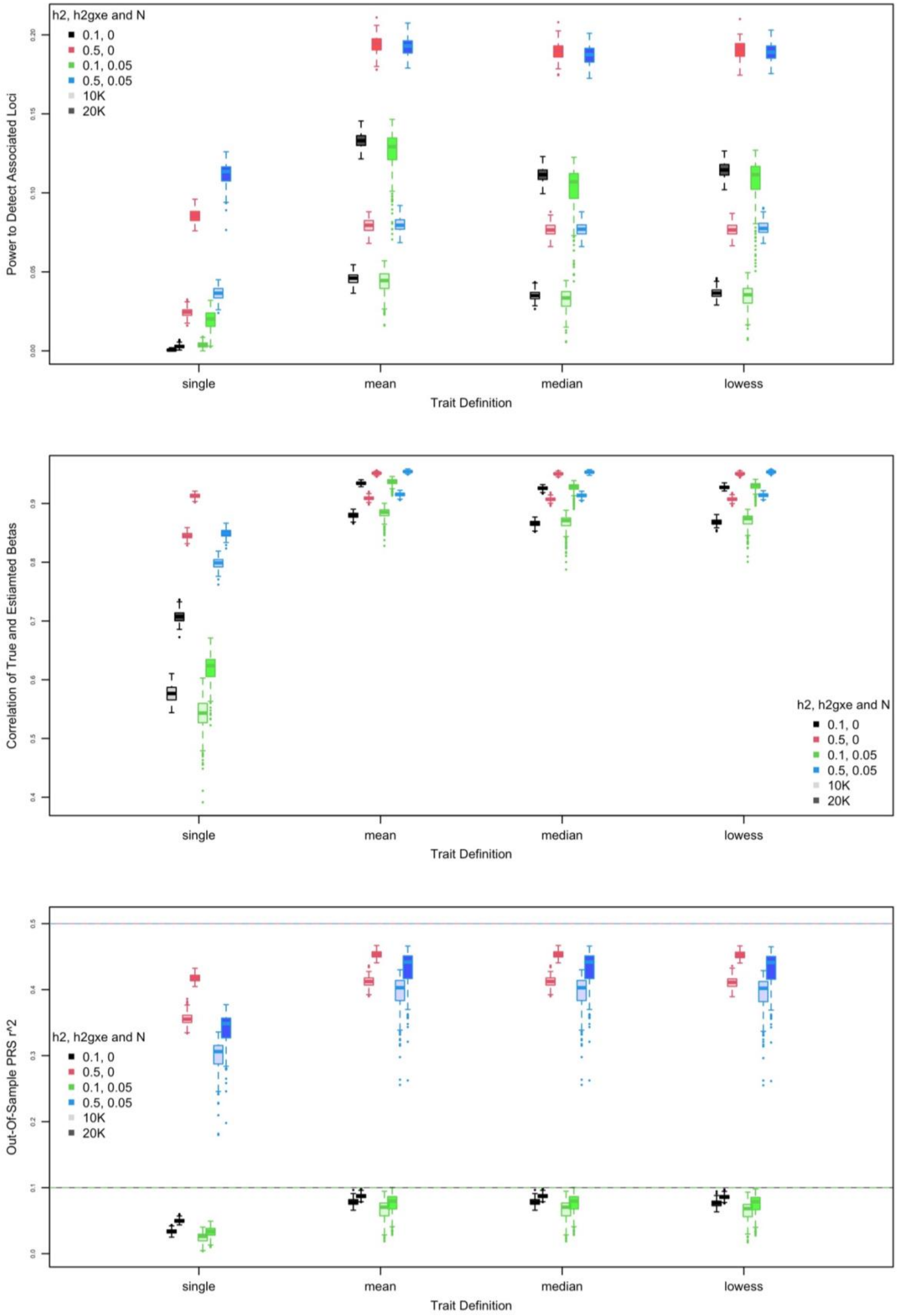


**Fig. S4.** Effect of discovery sample size (N) on simulated power, true-estimated beta correlation, and prediction R2. h2 and h2gxe represent the simulated heritability and simulated gene-by-time variance as a proportion of overall phenotypic variance, respectively.

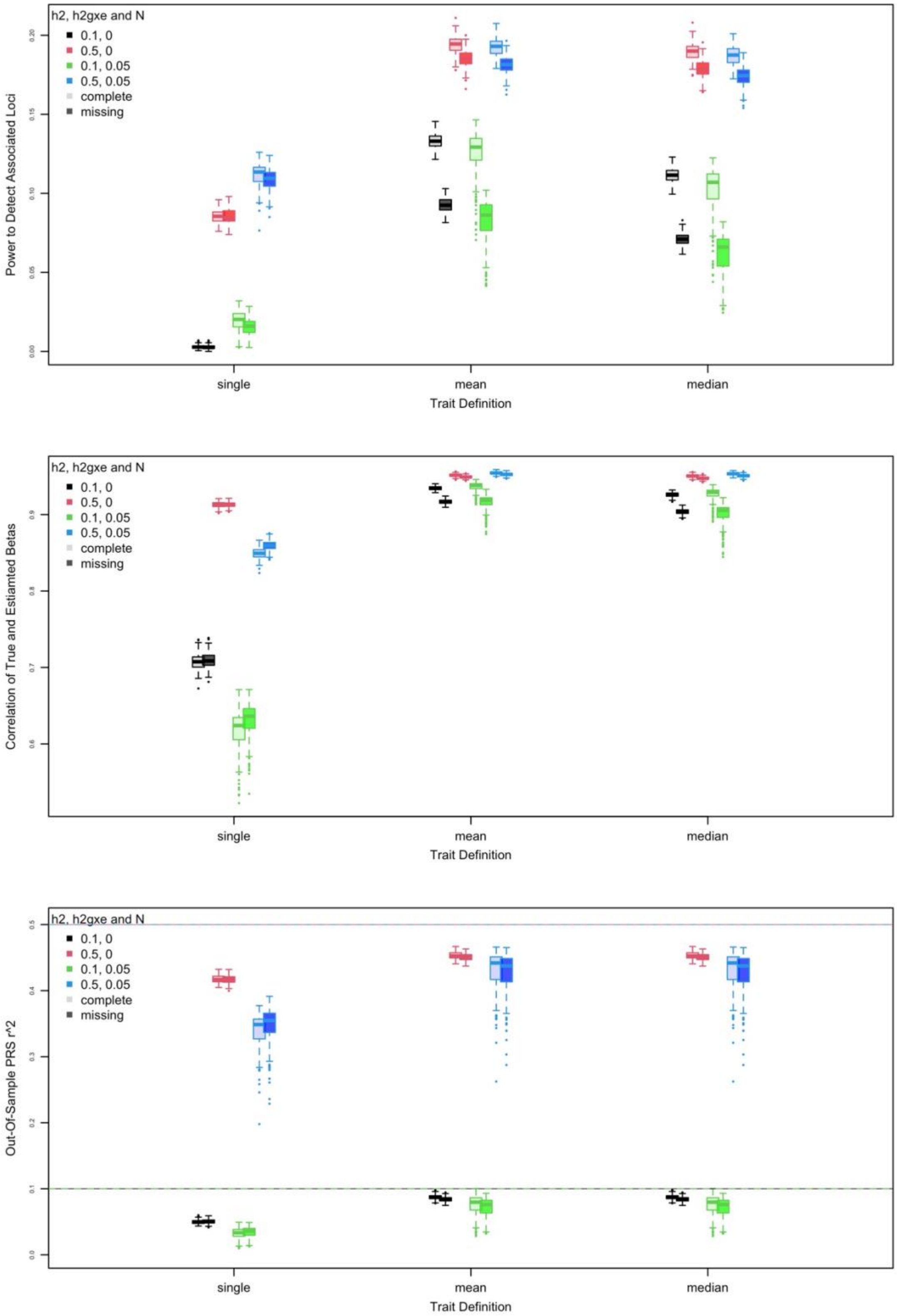


**Fig. S5.** Effect of discovery sample missingness (N) on simulated power, true-estimated beta correlation, and prediction R2. h2 and h2gxe represent the simulated heritability and simulated gene-by-time variance as a proportion of overall phenotypic variance, respectively. Here, "missing" corresponds to all individuals missing half the wave observations, chosen at random, compared to "complete" records in which all waves had observations for all individuals. Note that lowess curves could not be reliably fit when so much missingness was in the data, and we exclude those comparisons here.
